# Prenatal multiple micronutrient-fortified balanced energy-protein supplementation and newborn telomere length and mitochondrial DNA content: a randomized controlled efficacy trial in rural Burkina Faso

**DOI:** 10.1101/2023.11.22.23298825

**Authors:** Giles T. Hanley-Cook, Yuri Bastos-Moreira, Dries S. Martens, Trenton Dailey-Chwalibóg, Laeticia Celine Toe, Brenda de Kok, Lionel Ouédraogo, Alemayehu Argaw, Kokeb Tesfamariam, Patrick Kolsteren, Lieven Huybregts, Tim S. Nawrot, Sarah De Saeger, Marthe De Boevre, Carl Lachat

## Abstract

**Background:** Evidence regarding the effectiveness of prenatal nutritional supplements has mainly considered anthropometric pregnancy outcomes. The effect on markers of health and disease, such as offspring telomere length (TL) and mitochondrial DNA content (mtDNAc) is unknown.

**Objectives:** We assessed the efficacy of maternal multiple micronutrient (MMN)-fortified balanced-energy protein (BEP) and iron-folic acid (IFA) supplementation on newborn TL as a secondary outcome and mtDNAc as a non-declared outcome.

**Design:** We conducted a randomized controlled trial in rural Burkina Faso, among pregnant females (15-40 years old) enrolled at <21 weeks of gestation. Mothers received either MMN-fortified BEP and IFA (intervention) or IFA only (control) throughout pregnancy. Whole arterial blood samples were collected from the umbilical cord of 104 control and 90 intervention group infants, respectively. Average relative TL and mtDNAc were measured using quantitative polymerase chain reaction. Linear regression models were fitted to assess TL and mtDNAc differences across trial arms.

**Results:** We found that a combined daily MMN-fortified BEP supplement and IFA tablet did not affect newborn TL [β = -0.010 (95% CI: -0.057, 0.036); *P* = 0.662] or mtDNAc [β = 0.065 (95% CI: -0.203, 0.073); *P* = 0.354], as compared to an IFA tablet alone. These findings were confirmed (*P* >0.05) by adjusting the regression models for potential prognostic factors of study outcomes at enrollment. Exploratory analyses indicated higher, but non-significantly different mtDNAc among children born either small-for-gestational age, low birthweight, or preterm.

**Conclusions:** Newborns from mothers who received daily nutritional supplements across gestation did not have different relative TL or mtDNAc.

## Introduction

Prenatal multiple micronutrient (MMN) and balanced-energy protein (BEP) supplementation are strategies proposed to tackle maternal nutrient deficiencies and consequently reduce risks of small-for-gestational age (SGA), low birth weight (LBW), stillbirth, and increased birth weight, among malnourished females (1). Evaluating the effects of prenatal nutritional interventions on newborn biomarkers, rather than only on anthropometric outcomes (e.g., SGA, LBW), might provide more granular insights into nutritional impacts *in utero*. Moreover, such assessments likely further advance the mechanistic understanding of the developmental origins of health and disease phenomenon and subsequently help predict long-term risks, including disease vulnerability. Two potential biomarkers of interest are telomere length (TL) (2) and mitochondrial DNA content (mtDNAc) at birth (3).

The developmental origins of health and disease hypothesizes that harmful intrauterine exposures, including suboptimal maternal nutrition (4), might program the fetus to develop chronic diseases and overweight and obesity, when in a more favorable nutritional environments later in life (5). The presumed cellular adaptations are thought to involve epigenetic modifications, which amend their life-long expression without altering deoxyribonucleic acid (DNA) sequences (6–8).

Telomeres are non-coding, double-stranded, tandem repeats [5’-(TTAGGG)*_n_*-3’] of nucleotide sequences located at the tip of chromosomes. Telomeres maintain genome integrity, by protecting DNA coding sequences from degradation and preventing the aberrant fusion of chromosomes (9). In somatic cells, telomeres shorten after each cell division (i.e., 50-100 base pairs), due to incomplete replication of DNA molecules and maintenance mechanisms that are not able to prevent telomere attrition (10). TL shortening results in DNA damage, interruption of cellular function, chromosomal fusion, and cell senescence. TL at birth is a biological marker that may shape the human aging-phenotype later in life (11). Shorter TL has been consistently associated with higher mortality (12) and chronic disease rates, such as cardiovascular disease (13) and type II diabetes (14).

Mitochondria are known for their role in energy production by aerobic respiration in the form of adenosine triphosphate (ATP) (15). Predominantly maternally inherited, circular, double-stranded mtDNA is in theory more prone to damage due to oxidative stress-induced damage, caused by its proximity to the sites of oxidative phosphorylation (e.g., reactive oxygen species), and lack of protection from histones present in nuclear DNA (16,17). In pregnancies ending in fetal growth restriction or LBW, mtDNA has been observed to be elevated in maternal blood (18) and placental samples (19), indicating mitochondrial dysfunction. In the neonatal period and during infancy, mitochondrial dysfunction has been related to a myriad of clinical symptoms, including cerebral ataxia, poor weight gain, and heart arrhythmia (20,21), and the pathogenesis of chronic diseases in adulthood, including Alzheimer’s and cancer (22).

To date, two experimental studies have assessed the effectiveness of MMN-fortified supplements on children’s TL. In Ghana, prenatal supplementation had no impact on TL at 4-6 years of age as compared to iron-folic acid (IFA) (23), while in Bangladesh combining improved water, sanitation, and hygiene practices with child micronutrient fortified lipid-based nutrient supplementation (6-24 month of age) led to shorter TLs at 1 year, indicating increased telomere attrition, as compared to the control group (24). Furthermore, prenatal ω-3 supplementation did not increase offspring TL in Australia (25). Likewise, no beneficial effects were observed on TL in adults during a 5-year Mediterranean diet or a 12-week almond-enriched dietary interventions in Spain (26) and Australia (27), respectively. However, adults receiving a daily combination of vitamin supplements for 6 to 12 months had longer TLs in Greece, as compared to the control group (28). In addition, one trial assessed the impact of prenatal MMN supplementation on females’ mtDNA in maternal venous blood prior to delivery in Indonesia, and reported lower post-supplementation mtDNA compared to IFA, indicating improved mitochondrial efficiency (29).

The MISAME-III randomized controlled trial (RCT) assessed the efficacy of a prenatal MMN-fortified BEP supplement and IFA tablet among pregnant females in rural Burkina Faso, as compared to IFA alone on newborn relative TL and mtDNAc (30). Newborn TL was registered as a secondary outcome but mtDNAc was as non-declared outcome that was considered relevant for the trial during the analysis of the samples. The findings from these analyses will help characterize the physiological impact of MMN-fortified BEP supplementation, given the previously reported modest effects on the primary outcomes at birth and linear growth of infants at 6 months (31).

## Methods

Our research was reported using the Consolidated Standards of Reporting Trials (CONSORT) 2010 checklist (32) and Minimum Information for Publication of Quantitative Real-Time PCR Experiments (MIQE) guidelines (33).

### Study setting

The prenatal phase of the MISAME-III study was took place between the first enrolment on 30 October 2019 and the last delivery on 7 August 2021 in the catchment areas of six rural health centers of the health district of Houndé, Tuy Province, in the Hauts-Bassins region of Burkina Faso (34,35). Malaria transmission is perennial, with seasonal variations. The usual diet during pregnancy is non-diverse (36), predominantly maize-based with a complement of leafy vegetables (37), and consequently dietary micronutrient intakes are inadequate to cover the Estimated Average Requirements (EARs) (38). Moreover, among a subsample of MISAME-III females the mean energy intake of the base diet (i.e., excluding supplements) was estimated to be ∼1,940 kcal in both trial arms at the end of the pre-harvest season (39).

### Study design, participants, and enrolment procedures

The MISAME-III protocol containing all procedures was published previously (30). The study was a community-based, non-blinded individually randomized 2 × 2 factorial RCT, with directly observed daily supplement intake. The RCT specified one primary prenatal outcome: SGA [<10^th^ percentile of the International Fetal and Newborn Growth Consortium for the 21st Century (INTERGROWTH-21^st^) new-born size standards (40)]. Secondary and exploratory biological outcomes of the prenatal BEP intervention were relative newborn TL and mtDNAc at birth, respectively.

Females aged between 15 and 40 years and living in the study villages were identified by way of a census in the study area (*n* = 10,165). A network of 142 locally trained community support staff visited all eligible females at their homes every five weeks to identify pregnancy early, by screening for self-reported amenorrhea. Females suspected to be pregnant were referred to the health center for a urine pregnancy test. Once gestation was preliminarily confirmed, the MISAME-III study purpose and procedures were explained in the local language: Bwamu, Mooré, or Dioula. Prior to randomization, we excluded females who intended to leave the study area during their pregnancy, planned to deliver outside the study area, or mothers who had a peanut allergy since the BEP supplement is an energy-dense peanut paste fortified with MMNs (31).

After written informed consent was obtained, females (pregnancy not yet confirmed by an ultrasound) were randomly assigned to receive either a daily MMN-fortified BEP supplement and IFA tablet (intervention group) or a daily IFA tablet alone (control group) during pregnancy.

The stratified randomization scheme per health center was generated by an external research analyst before the start of the study with Stata 15.1 (StataCorp, College Station, TX), in permuted blocks of eight (4 control, 4 intervention). The allocation was coded with the letter A for the prenatal control arm and the letter B for the prenatal intervention arm. Randomization codes were concealed in sequentially numbered sealed opaque envelopes by project employees, who were not in direct contact with enrolled women. The project midwives who enrolled participants, assigned women to a trial arm by drawing a sealed envelope containing the A/B letter code. MISAME-III enrolment ran from 30 October 2019 to 12 December 2020. Within 14 days of enrolment, a female’s pregnancy was definitively confirmed by an ultrasound. Gestational age (GA) was estimated by measuring crown-rump length (7-13 weeks) or by calculating the mean of three to four measurements: bi-parietal diameter, head circumference, abdominal circumference, and femur length (12-26 weeks) (23). Post-randomization, we excluded non-pregnant women, mothers with a GA ≥21 completed weeks, and multi-fetal pregnancies (i.e., not meeting the *a priori* defined study inclusion criteria) (41).

Trained village-based project workers visited 10-25 pregnant females per day to ensure the directly observed intake of MMN-fortified BEP supplements and IFA tablets. When females had a short and scheduled absence from home, MMN-fortified BEP and IFA were given to the pregnant females in advance (thus, counted as non-observed intakes for the respective days). The home visitors also encouraged pregnant females to attend at least four scheduled antenatal care (ANC) visits approximately every seven weeks. All serious adverse events (e.g., miscarriage and stillbirth) were recorded on a case-by-case basis, and verbal autopsies were performed by the MISAME-III physician for maternal or infant deaths that occurred outside a health center.

Newborn arterial umbilical cord blood samples were collected from April 2021 onwards, at which time 304 females (164 control, 140 intervention) were still actively enrolled in the MISAME-III trial.

### Study supplements

In 2016, the Bill & Melinda Gates Foundation convened an expert group to recommend the optimal nutritional composition of the BEP supplement (42). In a formative study, the most preferred and suitable MMN-fortified BEP supplement was selected for administration in the MISAME-III efficacy trial (43,44). The BEP supplement is a lipid-based nutrient supplement in the form of an energy-dense peanut paste fortified with MMN. The BEP is ready-to-consume, does not require a cold chain, and has a long shelf life. On average, the 72g MMN-fortified BEP provided 393 kcal and consisted of 36%, 20%, and 32% energy from lipids, protein, and carbohydrates, respectively. Furthermore, the MMN content alone covered at least the daily EARs of micronutrients for pregnant females, except for calcium, phosphorous, and magnesium (45). The complete nutritional composition of the MMN-fortified BEP is provided in **Supplemental Table 1**.

Females in the intervention group received a daily MMN-fortified BEP supplement and an IFA tablet [65 mg iron (form: FeH_2_O_5_S) and 400 μg folic acid (form: C_19_H_19_N_7_O_6_; Tolerable Upper Intake Level from fortified food or supplements, not including folate from food: 1000 μg/d (46))], whereas females in the control group received a daily IFA tablet only (Sidhaant Life Sciences, Delhi, India), in accordance with Burkina Faso’s national health protocol (i.e., standard of care). Following Burkinabe guidelines, all enrolled females received malaria prophylaxis (three oral doses of sulfadoxine-pyrimethamine) at the relevant ANC visits.

### Data collection and measures

At enrolment (i.e., first ANC visit), we measured maternal height, weight, mid-upper arm circumference (MUAC) in duplicate, and hemoglobin (Hb) concentration. In addition, a comprehensive socio-economic and demographic questionnaire was administered at baseline (30).

Maternal height was measured to the nearest 1 cm using a ShorrBoard® Infant/Child/Adult (Weigh and Measure, Olney, MD) and weight to the nearest 100 g with a Seca 876 scale (Seca, Hanover, MD); and the accuracy of the scales was verified weekly. Maternal MUAC was measured to the nearest 1 mm using a Seca 212 measuring tape (Seca, Hanover, MD). Pregnant females’ Hb concentration was measured by spectrophotometry with a HemoCue® Hb 201+ (HemoCue, Ängelholm, Sweden); and a weekly calibration check was made with the use of a HemoCue Control Cuvette. The study’s physician performed trans-abdominal ultrasound fetal biometry within two weeks of enrollment. Pregnancy was confirmed and GA was estimated using a portable diagnostic imaging and full-color, flow-mapping SonoSite M-Turbo (FUJIFILM SonoSite Inc., Bothell, WA). Concurrently, maternal subscapular and tricipital skinfold measurements were taken in triplicate using a Harpenden caliper.

At birth, anthropometry of all newborns was assessed in duplicate within 12 hours by study midwives at the health center. Birth weight was measured to the nearest 10 g with a Seca 384 scale (Seca, Hanover, MD). If there was a large discrepancy between weight measures (i.e., >200 g), a third measurement was taken. The average of the two closest measures were used for analyses (i.e., SGA and LBW). The accuracy and precision of anthropometric measurements were established regularly through standardization sessions organized by an expert in anthropometry (47).

The arterial umbilical cord blood collection procedure was described in detail previously (48). Within 30 minutes post-partum, arterial umbilical cord blood (*n* = 195) was sampled, by an arterial puncture, into 4 mL BD Vacutainer® plastic whole blood tubes with spray-coated K2 potassium salt of ethylene diamine tetra acetic acid (EDTA) (BD, Franklin Lakes, NJ), which was gently inverted (at least 10 times) for thorough mixing of blood with the anticoagulant. Blood samples were aliquoted, using micropipettes (Thermo Fisher Scientific, Life Technologies, Europe), into sterile cryotubes (Biosigma, Cona VE, Italy) and flash frozen in 12 L liquid nitrogen storage vessels (Cryopal, Air Liquide, Paris, France). Umbilical cord blood samples collected after working hours (i.e., after 18:00 until 06:00) were gently inverted to mix and placed at 0-4 °C, before being aliquoted and flash frozen. Once liquid nitrogen storage vessels were full, samples were transferred to a -80 °C freezer at the Institut de Recherche en Sciences de la Santé, Bobo-Dioulasso, Burkina Faso. Samples were shipped in dry ice to the allocated biobank of the MISAME-III study in the Centre of Excellence in Mycotoxicology and Public Health, Faculty of Pharmaceutical Sciences, Ghent University and stored at -80 °C. For DNA extraction, samples were transferred in dry ice to the Centre for Environmental Sciences, Hasselt University and stored at -80°C. All blood samples were subjected to one freeze-thaw cycle.

MISAME-III field data were collected using SurveySolutions v.21.5 on tablets by the project physician and midwives, which were transferred to a central Ghent University server weekly. Questionnaire assignments were sent once a week to the field team including preloaded data collected at a previous ANC visit to lower the amount of incorrect data. Furthermore, we programmed generic validation codes to avoid the entry of implausible values and to improve the quality of data collection in the field. Additionally, bi-weekly data quality checks and missing or inconsistent data were sent back to the field for revision. The quality of ultrasound images and estimation of GA were checked for >10% of the examinations regularly by an external gynecologist, using a quality checklist and scoring sheet. The trained project workers collected daily data on MMN-fortified BEP and IFA compliance in both prenatal study arms by smartphone-assisted personal interviewing programmed in CSPro v.7.3.1. Six supervisors performed monthly Lot Quality Assurance Sampling schemes of each home visitor’s work on a random day (49).

### Relative telomere length and mitochondrial DNA content

DNA extraction was performed using the QIAamp DNA Mini Kit (Qiagen, Inc., Venlo, the Netherlands). DNA quantity and purity were assessed by a Nanodrop 1000 spectrophotometer (Isogen, Life Science, Belgium). DNA was considered pure when the A260/280 was greater than 1.80 and A260/230 greater than 2.0. All isolated DNA was stored at -25 °C at the molecular biology laboratory of the Centre for Environmental Sciences, Hasselt University.

DNA integrity was assessed by agarose gel-electrophoresis. To ensure a uniform DNA input of 5 ng for each quantitative polymerase chain reaction (qPCR), samples were diluted and checked using the Qubit 1X dsDNA HS Assay Kit (Thermo Fisher Scientific, Life Technologies, Europe) (50).

Relative TL and mtDNAc were measured using a real-time qPCR method in which telomere specific amplification is performed using primers described by Cawthon (51), as well as mtDNA amplification using primers (i.e., targeting the ND1 mitochondrial gene) described by Janssen *et al.* (52). A single-copy gene (S), in this case, human beta globin specific primers were used (53). The telomere-specific qPCR reaction mixture contained 1x KAPA SYBR Fast PCR master mix (Merck KGaA, Darmstadt, Germany), 2 mM dithiothreitol (DTT), 100 nM telg primer (5’-ACACTAAGGTTTGGGTTTGGGTTTGGGTTTGGGTTAGTGT-3’) and 100 nM telc primer (5’-TGTTAGGTATCCCTATCCCTATCCCTATCCCTATCCCTAACA- 3’). The following cycling conditions were applied: 1 cycle at 95 °C for 3 minutes, 2 cycles at 94 °C for 3 seconds, 49 °C for 15 seconds, 30 cycles at 94 °C for 3 seconds, 62 °C for 5 seconds, and 74 °C for 10 seconds. The mtDNA (MT-ND1) reaction mixture contained 1x KAPA SYBR Fast PCR master mix, 450 nM forward (5’- ATGGCCAACCTCCTACTCCT- 3’), 450nM reverse (5’- CTACAACGTTGGGGCCTTT-3’) primer, and 5 ng DNA. The cycling conditions were: 1 cycle at 95 °C for 3 minutes, 40 cycles at 95 °C for 3 seconds, and 58 °C for 15 seconds. The single-copy gene qPCR mixture contained 1x KAPA SYBR Fast PCR master mix, 450 nM HBG1 primer (5’-GCTTCTGACACAACTGTGTTCACTAGC-3’), and 450 nM HBG2 primer (5’- CACCAACTTCATCCACGTTCACC-3’). Cycling conditions were similar for mtDNA. All measurements were performed in triplicate on a QS5 Fast Real-Time PCR System (Applied Biosystems, Waltham, MA) in a 384-well format (54).

After each qPCR a melting curve analysis was performed. On each run, PCR efficiency was evaluated using two standard 6-point serial diluted standard curves (108% for TL, 120% for MT-ND1, and 96% for SCG with an *R^2^* >0.994 for all standard curves). Nine inter-run calibrators (IRCs) were run to account for inter-run variability. Relative leukocyte TL and mtDNAc were calculated using the qBase software (Biogazelle, Zwijnaarde, Belgium). In qBase, TL and mtDNA are calculated as a calibrated normalized relative quantity (CNRQ) (55). The latter is achieved by first calculating the relative quantity (RQ) based on the delta-Cq method for telomere copy number (T), mitochondrial gene copy number (M), and S obtained Cq values, using target specific amplification efficiencies. Since the choice of a calibrator sample (i.e., sample to which subsequent normalization is performed, delta-delta-Cq) strongly influences the error on the final RQs (due to measurement errors on the calibrator sample), normalization is performed to the arithmetic mean quantification values for all analyzed samples, which results in the normalized relative quantity (NRQ). Finally, as samples are measured over multiple qPCR plates, 9 IRCs are used to calculate an additional correction factor to eliminate run-to-run differences, resulting into the final T/S and M/S ratios (i.e., CNRQ). Mathematical calculation formulas to obtain RQ, NRQ, and CNRQs are provided by Hellemans *et al*. (55).

The inter- and intra-assay repeatability were assessed by calculating the intraclass correlation coefficient (ICC) with 95% CI of triplicate measures for the IRCs (inter-assay) and all T/S ratios and M/S ratios (intra-assay) the ICC R-code provided by the Telomere Research Network (56). The inter-assay ICC for TL was 0.97 (95% CI: 0.88, 0.99; *P* <0.0001) and 0.93 (95% CI: 0.74, 0.98; *P* <0.0001) for mtDNAc. The intra-assay repeatability for TL and mtDNAc were 0.89 (95% CI: 0.86, 0.91; *P* <0.0001) and 0.96 (95% CI: 0.95, 0.97; *P*<0.0001).

### Statistical analysis

For consistency and comparability of study findings, the present complete cases analyses followed the analytical procedures used to assess the efficacy of the prenatal MMN-fortified BEP intervention on birth (31) and maternal outcomes (57). The MISAME-III statistical analysis plan was validated on October 24, 2019 and published online on November 3, 2020 on the study’s website: https://www.misame3.ugent.be/resource-files/MISAME-III_SAP_v1_102019.pdf.

In line with the analysis of primary outcomes, we restricted our analyses to singleton pregnancies. Descriptive data are presented as frequencies (%), means ± SDs, or medians [interquartile ranges (IQRs)]. Unadjusted and adjusted group differences were estimated by fitting linear regression models for the continuous T/S and M/S outcomes, to estimate the mean group difference. The assumptions of normality were checked by visual inspection of the standardized normal probability and quantile-quantile plots of the residuals (**Supplemental Figure 1**, **Supplemental Figure 2**). All models were adjusted for health center, randomization block, and qPCR plate as a fixed effects to account for clustering by the study design. Adjusted models additionally contained a set of potential baseline prognostic factors of newborn TL and mtDNAc, including wealth index (0-10 points), maternal age (years), primiparity, GA (weeks), height (cm), MUAC (mm), body mass index (kg/m²), and Hb level (g/dL) at study enrolment. We did not adjust for any other socio-demographic variables, due to balanced baseline characteristics across prenatal study groups (i.e., <|2.5| percentage points difference). As sensitivity analyses, we additionally included the age (years) of the household head at enrolment as a proxy for the father’s age (58) and the time (minutes) between umbilical cord blood collection and storage in the liquid nitrogen vessel.

We followed an approach used by Katz *et al.* (59), Roberfroid *et al.* (60), and de Kok *et al.* (31) to explore whether the treatment effects on T/S and M/S ratios were constant over percentiles of newborns’ T/S and M/S ratios distributions, respectively. In this method, differences (and CIs) in biological outcomes between intervention and control groups were estimated as nonlinear smooth functions of every aggregated 4^th^ percentile of newborn TL or mtDNAc distributions, with knots set at the 20^th^, 40^th^, 60^th^, and 80^th^ percentiles. Furthermore, we conducted stratified analyses by child sex, to evaluate any potential effect modification. Lastly, as an exploratory analysis, we assessed TL and mtDNAc across adverse birth phenotypes (i.e., SGA, LBW, or preterm).

Statistical significance was set at *P* <0.05 for all two-sided tests, except for exploratory interactions for which *P* <0.10 was used. All analyses were conducted with Stata 16.1 (StataCorp, College Station, TX).

## Results

Between October 2019 and December 2020, 2,016 females were assessed for eligibility, of whom 1,897 were randomized (960 control, 937 intervention) and 119 excluded for not meeting the MISAME-III’s inclusion criteria. In April 2021, 304 females (164 control, 140 intervention) were still participating in the efficacy trial. Among the 195 pregnant females, from whom umbilical cord blood samples were collected between April 2021 and August 2021 (105 control, 90 intervention), one female was excluded from the control group post-randomization, due to her GA at inclusion being ≥21 completed weeks (i.e., confirmed by an ultrasound, after an initial urine pregnancy test). Lastly, two (0 control, 2 intervention) and three women (1 control, 2 intervention) were removed from T/S and M/S ratio analyses, due to insufficient DNA (**Figure 1**). The median (IQR) time between umbilical cord blood collection and storage in the liquid nitrogen vessel was 0 (0, 491) minutes in the control group and 0 (0, 550) minutes in the intervention group.

**Figure 1.**
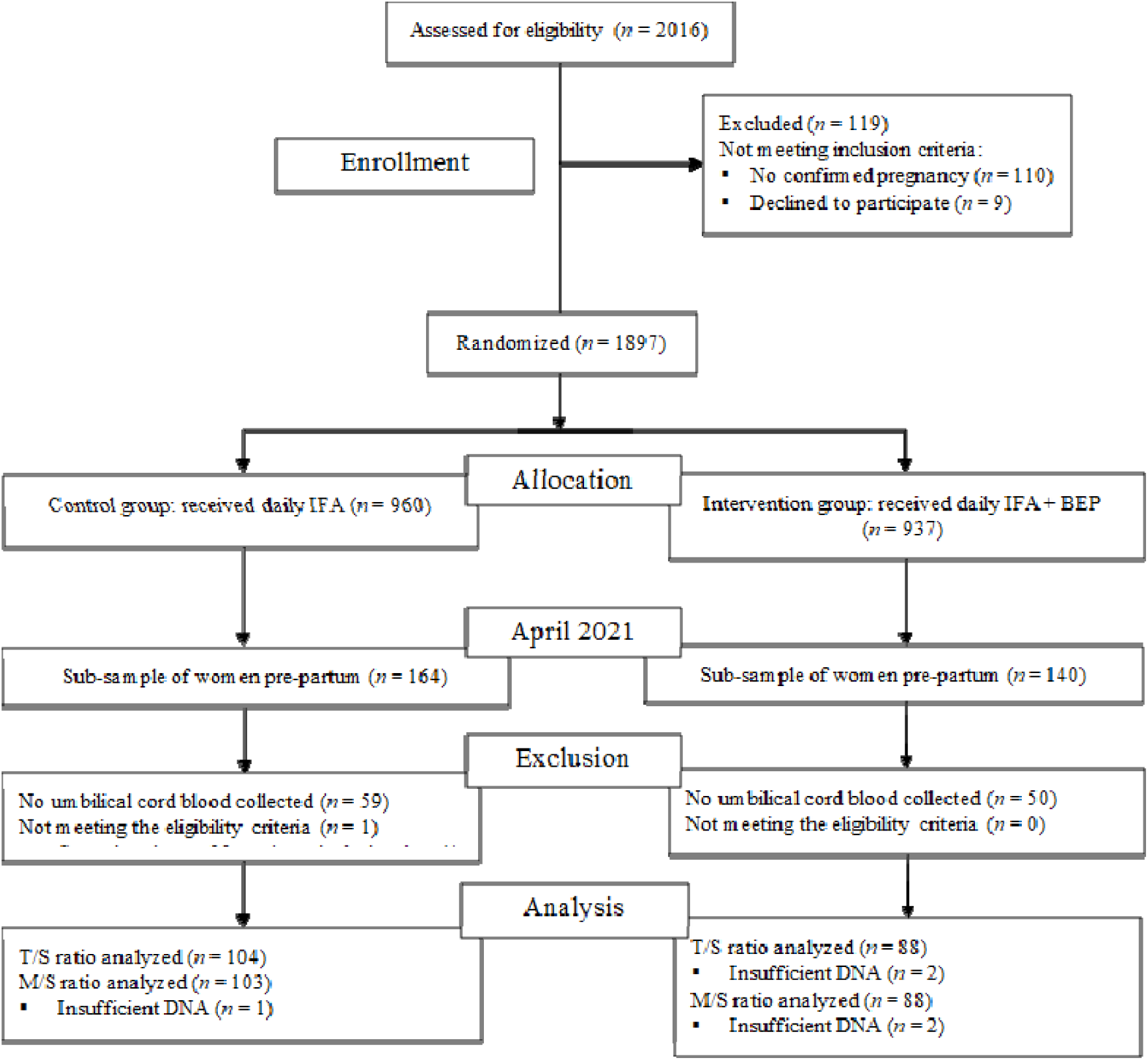
MISAME-III trial flow chart. DNA, deoxyribonucleic acid; T/S, telomere copy number to a single-copy gene number; MISAME-III, Micronutriments pour la Santé de la Mère et de l’Enfant study 3; M/S, mitochondrial gene copy number to a single-copy gene number.

The baseline characteristics of eligible pregnant females (104 control, 90 intervention) are presented in **Table 1**. In response to a reviewer comment regarding potential baseline imbalances, and in recognition that baseline statistical tests in randomized controlled trials are not fundamentally incorrect, *P*-values are provided in **Supplemental Table 2**. The prenatal trial arms were balanced regarding household, maternal, and pregnancy characteristics. At baseline, 70.1% of households were food insecure, 58.1% and 62.9% of households had improved water sources and sanitation, respectively, 46.4% of pregnant females completed primary education, 6.19% were underweight, 26.3% were anemic, and their mean ± SD GA was 9.59 ± 3.17 weeks. The prevalence of self-reported diabetes and oedema was 0% at inclusion. Moreover, the mean ± SD for systolic blood pressure in intervention and control groups were: 114 ± 3 and 113 ± 14 mmHg, respectively. On the other hand, diastolic blood pressure was 69 ± 9 mmHg for both trial arms. In our sub-study, the mean ± SD duration under supplementation was 28 ± 3 and 29 ± 3 weeks for intervention and control groups, respectively. Furthermore, the median (IQR) adherence to MMN-fortified BEP was 96% (78, 99) in the intervention group, while mean IFA compliance was >95% in both prenatal study arms.

**Table 1.** Baseline characteristics of study participants, by MISAME-III trial arm.

| Characteristics | Control (n = 104) | Intervention (n = 90) |
| --- | --- | --- |
| **Health centre catchment area** |  |  |
| Boni | 17 (16.3) | 16 (17.8) |
| Dohoun | 14 (13.5) | 13 (14.4) |
| Dougoumato II | 13 (12.5) | 14 (15.6) |
| Karaba | 11 (10.6) | 8 (8.89) |
| Kari | 21 (20.2) | 21 (23.3) |
| Koumbia | 28 (26.9) | 18 (20.0) |
| **Household level** |  |  |
| Wealth index, 0-10 points | 4.32 ± 1.72 | 5.02 ± 1.77 |
| Household food insecurity<sup>2</sup> | 72 (69.2) | 64 (71.1) |
| Improved primary water source<sup>3</sup> | 62 (59.6) | 52 (57.8) |
| Improved sanitation facility<sup>4</sup> | 68 (65.4) | 54 (60.0) |
| Household size | 5.85 ± 4.34 | 6.89 ± 4.74 |
| Polygamous households | 38 (36.5) | 40 (44.4) |
| **Head of household** |  |  |
| Age, years | 32.3 ± 8.83 | 32.2 ± 9.31 |
| Male | 103 (99.0) | 90 (100) |
| Completed primary education | 63 (60.6) | 55 (61.1) |
| **Maternal** |  |  |
| Age, years | 24.0 ± 5.47 | 24.0 ± 5.70 |
| **Ethnic group** |  |  |
| Bwaba | 57 (54.8) | 51 (56.7) |
| Mossi | 36 (34.6) | 30 (33.3) |
| Other | 11 (10.6) | 9 (10.0) |
| **Religion** |  |  |
| Muslim | 47 (45.2) | 41 (45.6) |
| Animist | 26 (25.0) | 22 (24.4) |
| Protestant | 23 (22.1) | 19 (21.1) |
| Catholic | 6 (5.77) | 6 (6.66) |
| No religion, no animist | 2 (1.92) | 2 (2.22) |
| Completed primary education | 53 (51.0) | 37 (41.1) |
| Weight, kg | 58.9 ± 10.7 | 58.1 ± 7.88 |
| Height, cm | 162 ± 5.83 | 163 ± 5.83 |
| BMI, kg/m<sup>2</sup> | 22.4 ± 3.54 | 21.9 ± 2.65 |
| <18.5 kg/m<sup>2</sup> | 8 (7.69) | 4 (4.44) |
| Mid-upper arm circumference, mm | 263 ± 31.9 | 261 ± 24.0 |
| Subscapular skinfold, mm | 12.5 ± 7.18 | 11.8 ± 5.55 |
| Tricipital skinfold, mm | 11.9 ± 5.53 | 11.5 ± 4.38 |
| Haemoglobin (Hb), g/dl | 11.8 ± 1.34 | 11.8 ± 1.39 |
| Anemia (Hb <11 g/dl) | 23 (22.1) | 28 (31.1) |
| Severe anemia (Hb <7g/dl) | 0 (0) | 0 (0) |
| Gestational age, weeks | 9.43 ± 2.96 | 9.78 ± 3.41 |
| Trimester of gestation |  |  |
| First | 86 (82.7) | 74 (82.2) |
| Second | 18 (17.3) | 16 (17.8) |
| Parity |  |  |
| 0 | 26 (25.0) | 26 (28.9) |
| 1-2 | 49 (47.1) | 32 (35.6) |
| ≥3 | 29 (27.9) | 32 (35.6) |

<sup>1</sup>Data are frequencies (%) or means ± standard deviation. BMI, body mass index; Hb, hemoglobin. MISAME-III, Micronutriments pour la Santé de la Mère et de l'Enfant study 3.
<sup>2</sup>Assessed using FANTA/USAID's Household Food Insecurity Access Scale (85). Missing for one female in the control group.
<sup>3</sup>Protected well, borehole, pipe or bottled water were considered improved water sources.
<sup>4</sup>Flush toilet connected to local sewage or septic tank, or pit latrine with slab and/or ventilation were considered improved sanitation facilities.

In the present study, the medians (IQRs) of T/S ratios were 1.00 (0.90, 1.12) in the control group and 1.00 (0.89, 1.11) in the intervention group, while M/S ratios were 1.11 (0.82, 1.31) and 1.00 (0.71, 1.33), respectively (**Figure 2**). Our unadjusted complete cases analyses of a combined daily MMN-fortified BEP supplement and IFA tablet indicated a non-significant difference in newborn T/S [-0.006 (95% CI: -0.052, 0.040); *P* = 0.810] or M/S ratios [-0.059 (95% CI: -0.196, 0.079); *P* = 0.403], as compared to an IFA tablet alone (**Table 2**). These findings were confirmed by adjusting the regression models for potential prognostic factors of study outcomes at enrollment (**Table 2**) and further adjustment for the age of the household head at inclusion and time between umbilical cord blood collection and storage in the liquid nitrogen vessel (data not shown).

**Figure 2.**
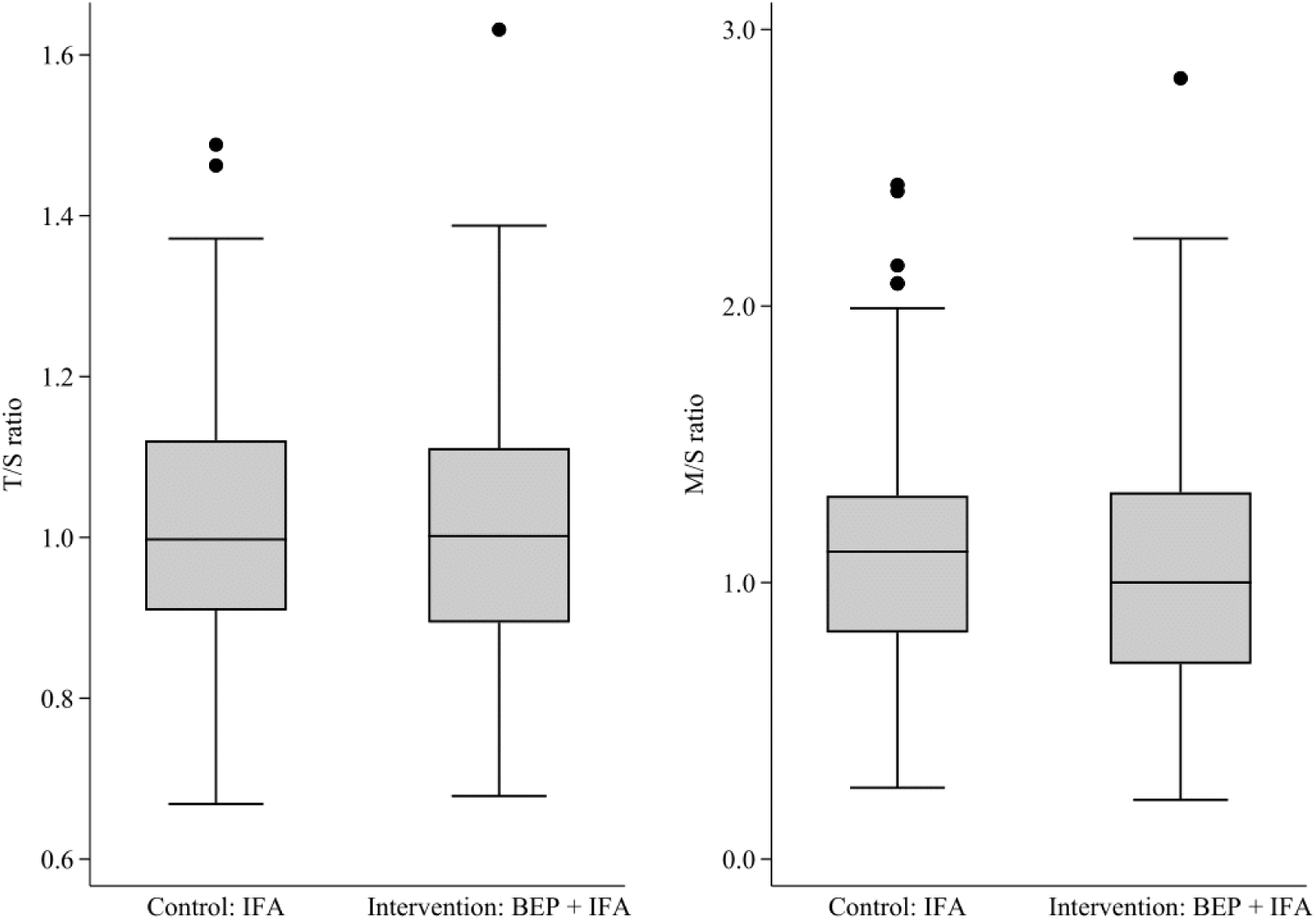
Box and whisker plots of newborn telomere length (left) and mitochondrial DNA content (right), by MISAME-III trial arm. T/S, telomere copy number to a single-copy gene number; MISAME-III, Micronutriments pour la Santé de la Mère et de l’Enfant study 3; M/S, mitochondrial gene copy number to a single-copy gene number.

**Table 2.** Efficacy of prenatal multiple micronutrie nt-fortified BEP supplementation on relative telomere and mitochondrial DNA content.

| **Birth characteristics** | **Control<sup>1</sup>**<br>**(n = 104)** | **Intervention<sup>1</sup>**<br>**(n = 88)** | **Unadjusted $\Delta^3$**<br>**(95% CI)** | **P-value** | **Adjusted $\Delta^3$**<br>**(95% CI)** | **P- value** |
| --- | --- | --- | --- | --- | --- | --- |
| T/S ratio | 1.01 ± 0.154 | 1.01 ± 0.163 | -0.006 (-0.052, 0.040) | 0.810 | -0.007 (-0.056, 0.042) | 0.773 |
| M/S ratio | 1.12 ± 0.438<sup>2</sup> | 1.05 ± 0.455 | -0.059 (-0.196, 0.079) | 0.403 | -0.047 (-0.194, 0.099) | 0.523 |

Furthermore, prenatal MMN-fortified BEP and IFA supplementation had an inconsistent effect on T/S ratio across the percentiles of the newborn TL distribution (**Figure 3**), while a stronger negative effect on M/S ratio was observed between the ∼20^th^ and ∼60^th^ percentile of the mtDNAc distribution (**Figure 4**). Moreover, (un)adjusted stratified analyses did not indicate an effective modification of the MMN-fortified BEP intervention on newborn TLs or mtDNAc by child sex (*P* _intervention × sex_ >0.10) (**Supplemental Table 3**). Lastly, descriptive analyses indicated higher, but non-significantly different M/S ratios among children born either SGA, LBW, or preterm (**Supplemental Table 4**).

**Figure 3.**
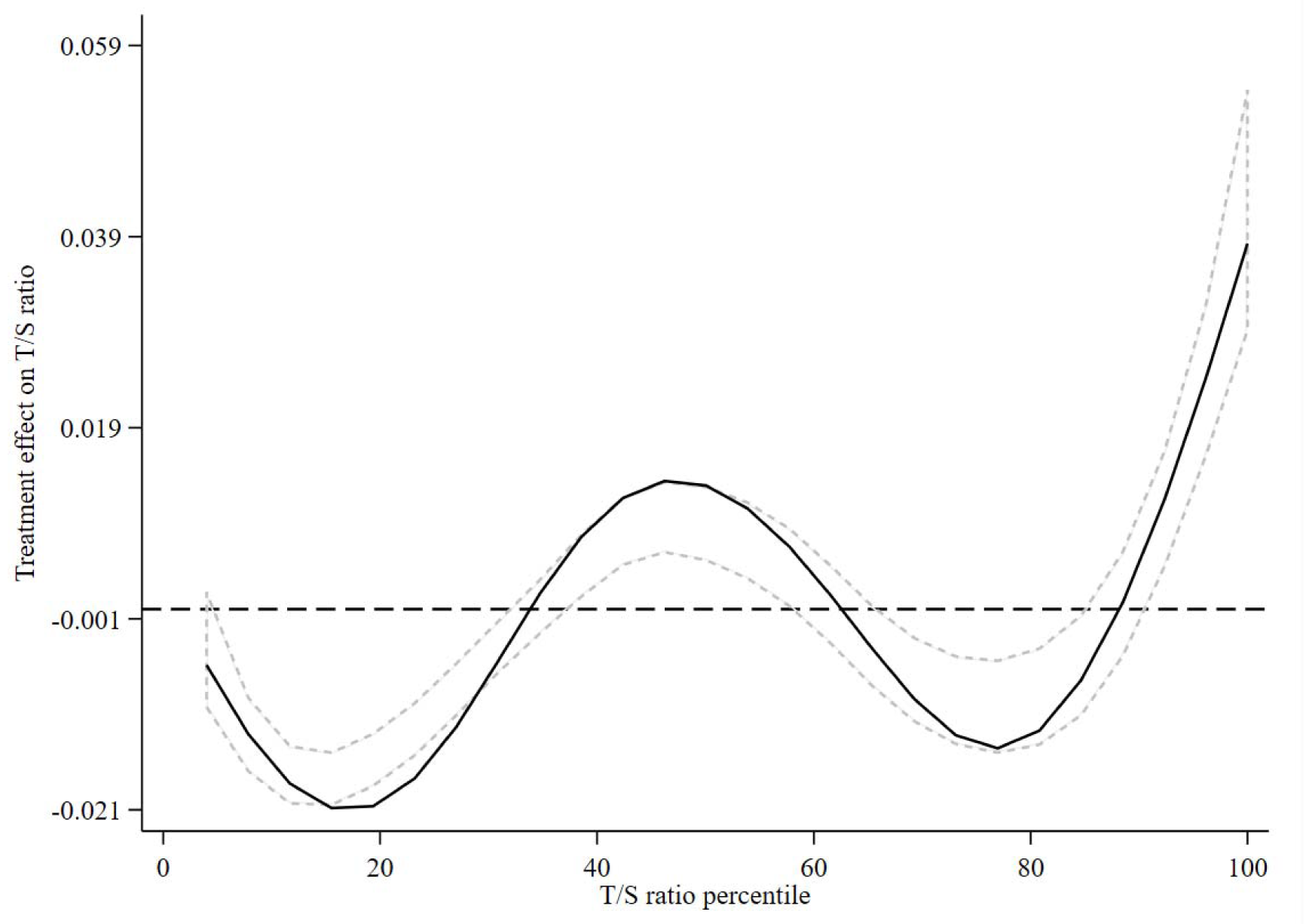
Treatment efficacy on T/S ratio across the distribution of T/S ratio. The estimated difference in T/S ratio between the women who received the MMN-fortified BEP supplement and IFA (intervention) and those who received only iron and folic acid (control) is shown as a function of the percentiles of T/S ratio. The zero line indicates no efficacy of the multiple micronutrient-fortified BEP. The positive y values indicate a higher T/S ratio in the intervention group, and the negative y values indicate a lower T/S ratio. The central solid black line represents the smoothed treatment efficacy, with upper and lower dashed 95% confidence bands, using complete cases. BEP, balanced energy-protein; IFA, iron-folic acid; MMN, multiple micronutrients; T/S, telomere copy number to a single-copy gene number.

**Figure 4.**
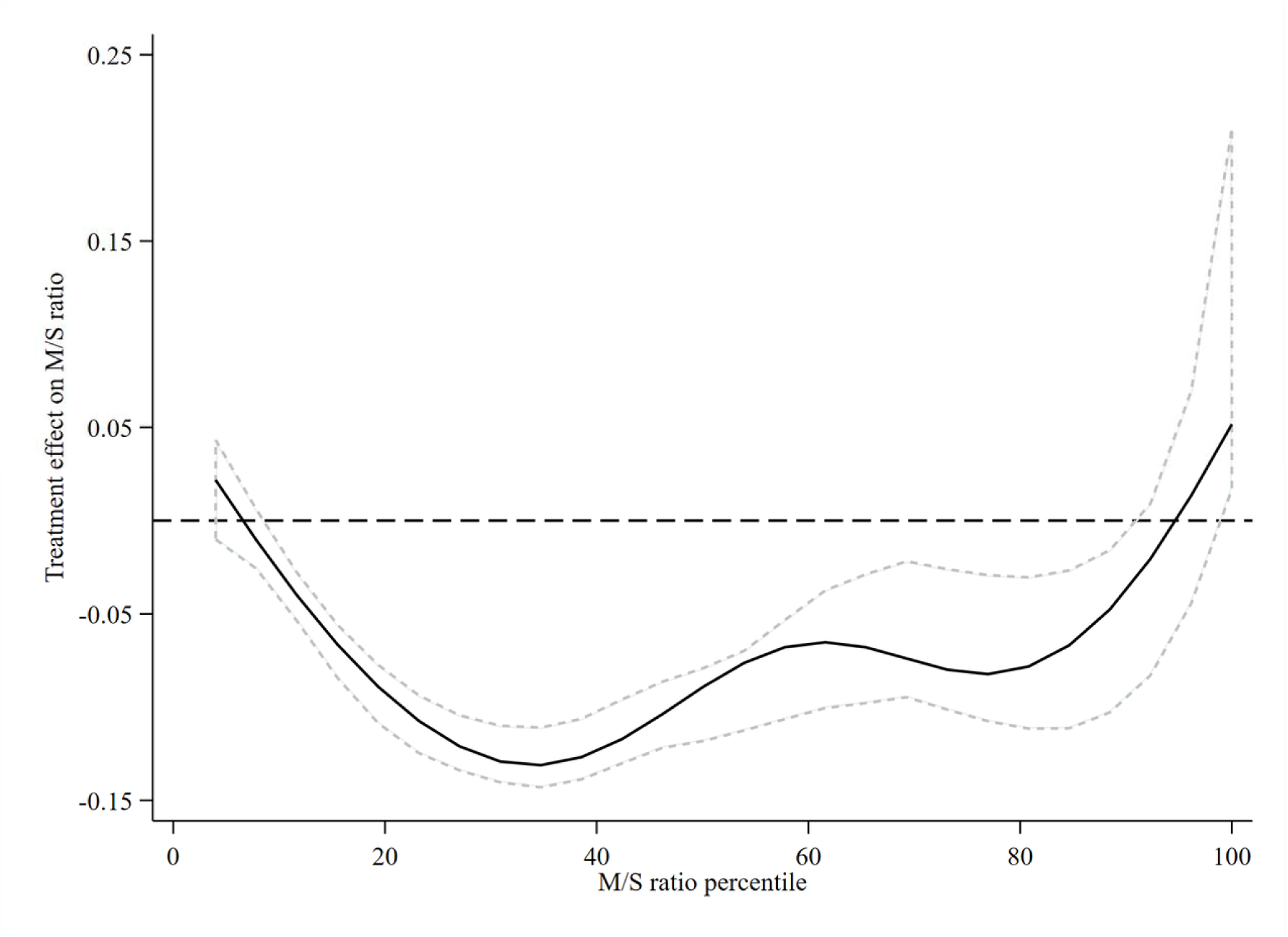
Treatment efficacy on M/S ratio across the distribution of M/S ratio. The estimated difference in M/S ratio between the women who received the MMN -fortified BEP supplement and IFA (intervention) and those who received only iron and folic acid (control) is shown as a function of the percentiles of M/S ratio. The zero line indicates no efficacy of BEP. The positive y values indicate a higher M/S ratio in the intervention group, and the negative y values indicate a lower M/S ratio. The central solid black line represents the smoothed treatment efficacy, with upper and lower dashed 95% confidence bands, using complete cases. BEP, balanced energy-protein; IFA, iron-folic acid; MMN, multiple micronutrients; M/S, mitochondrial gene copy number to a single-copy gene number.

## Discussion

In the MISAME-III trials, we found that newborns from mothers who received a daily MMN-fortified BEP supplement and IFA tablet did not have different relative TL or mtDNAc, as compared to the newborns of females who received an IFA tablet only. Nonetheless, offspring with adverse birth phenotypes (e.g., SGA, LBW) tended to have higher mtDNAc, likely indicating mitochondrial dysfunction.

Folate is a methyl donor and is a necessary precursor for fetal nucleotide synthesis and cell proliferation (61). However, in our trial, both the intervention and control groups received folate, so it is possible that IFA tablets alone had a saturated effect on TL that we were unable to observe due to the active control arm. Indeed, findings from a previous MISAME-III sub-study indicated that the nutrient requirements for folate were covered for all pregnant women (38). Nonetheless, it could be postulated that the vitamin B12 in BEP supplements might have reduced the amount of folate metabolically trapped as 5-methyltetrahydrofolate (62). The absence of an increase in TL at birth is in contrast with two prospective cohort studies, based in the United States of America and South Korea, indicating that higher maternal serum folate (61) and vitamin D concentrations during pregnancy are associated with longer TL in newborns (63). Moreover, vitamin D in the BEP upregulates telomerase activity, an essential enzyme for maintaining TL, while also promoting the expression of Klotho, a protein associated with longer TLs (64). Pregnancy is a state of low grade chronic inflammation, with the latter being associated with shorter TL (65). Poly-unsaturated fatty acids (e.g., ω-3) facilitate anti-inflammatory response, which was therefore hypothesized to lead to longer TL (23); nonetheless our null findings are consistent with a previous study that indicated prenatal ω-3 supplementation did not increase TL in Australia (25).The iron in the MMN-fortified BEP supplement, in combination with IFA, might have acted as a pro-oxidant (e.g., hydroxyl free radicals) and mitigated the anti-oxidant effects of other micronutrients (e.g., vitamin C and E) on TL (66). Similar to the current findings of prenatal macronutrient supplementation in MISAME-III (i.e., 393 kcal with <25% energy from protein), two prospective cohorts, in South Korea and the United States of America, did not find consistent associations between higher maternal protein, carbohydrate or fat intakes and their newborn’s TL (63,67).

The lack of impact of prenatal MMN-fortified BEP supplementation on newborn mtDNAc compare to findings in Cambodia, where daily iron supplementation (60 mg/day) led to a decrease in mtDNAc, but no changes in TL, after 12 weeks among non-pregnant females, as compared to placebo (68). Similarly, in Indonesia, prenatal MMN supplementation led to a lower maternal mtDNA copy number, as compared to IFA (29). Our MMN-fortified BEP supplement contained a plethora of vitamins (e.g., thiamin) and minerals, such as copper, iodine, selenium, and zinc, which was hypothesized to reduce oxidative stress, protect against mitochondrial damage and improve mitochondrial stability (69). Moreover, riboflavin is a key building block in mitochondrial complex I and II (70). The heterogeneity in study results might be explained by the dynamic nature of mitochondrial homeostasis, in which oxidative stress-induced damage to mtDNA (e.g., due to iron deficiency or overload) can lead to either mitochondrial biogenesis or mitophagy, and subsequently increased or decreased mtDNAc (71) This mitochondrial morphology adjustment is required to prevent metabolic insults, and consequently alters mitochondrial function to meet cellular energy and metabolic demands (72). Yet, findings from two American observational studies which indicated that higher adherence to healthy dietary patterns (73) and higher fruit and vegetables consumption (i.e., sources of antioxidants, such as vitamin C and E) are associated with greater mtDNA copy-numbers in adults (74).

In MISAME-III, we observed non-significant differences in newborn TL and mtDNAc across small samples of birth phenotypes (e.g., SGA *vs* non-SGA). Similarly, a meta-analysis did not find that intrauterine growth restriction was associated with shorter TL when measured in cord blood (75). In India, placental mtDNA copy number was significantly greater among SGA newborns (76), whereas umbilical cord blood mtDNAc was also significantly higher among growth restricted offspring and mothers who suffered preeclampsia in Italy (77). In parallel, higher maternal mtDNA copy number was associated with lower birth weights in Indonesia (18). Conversely, mtDNAc was lower in SGA and large-for-gestational age Argentinian newborns (3). However, fetal growth faltering was associated with shorter newborn TL when measured in placental tissue, while TL at birth was significantly longer in preterm birth than in full-term infants when measured by qPCR (75).

Our experimental study has several strengths. Compliance to BEP and IFA supplementation was verified by a community-based network of home visitors, resulting in high levels of observed adherence. Furthermore, quantitative 24-hour recalls confirmed that daily energy and micronutrients requirements were covered by consuming the MMN-fortified BEP supplement in combination with the usual diet and also ruled out any dietary substitution effects related to BEP supplementation (38). Moreover, considering that the umbilical cord blood collection procedure set up was delayed in the study, causing some pregnancies to be missed, and the remoteness of the study areas, where some participants gave birth outside of the health center or after working hours, there was still a high collection rate of samples. In addition, using qPCR, only small amounts of DNA were required (78); therefore, the assay was simple, rapid, and we were able to achieve a high throughput of the samples (53). Lastly, although there are alternative methods of telomere measurement, such as telomere restriction fragment and fluorescent in situ hybridization (FLOW-FISH), these techniques have limitations in population-based studies. Telomere restriction fragment requires large amounts of DNA; therefore, it is unsuitable for research with limited quantities of specimens. Likewise, FLOW-FISH requires fresh blood and so it is not applicable for samples stored at - 80°C (79).

Nonetheless, our study had some limitations that warrant caution. First, we did not assess females’ state of inflammation (e.g., C-reactive protein, fibrinogen, interleukin-6, tumor necrosis factor-alpha), stress (e.g., cortisol), or exposure to environmental contaminants (e.g., mycotoxins, black carbon) at inclusion, nor collect information on maternal physical activity and paternal age at enrolment, hence the use of the household head’s age as a proxy variable. Second, TL and mtDNAc were analyzed at birth only. Therefore, we are unable to evaluate the effect of MMN-fortified BEP supplementation on infants TL attrition and long-term changes in mitochondrial functioning. Third, due to limited sample volumes collected from the umbilical cord we were unable to obtain buffy coat from whole umbilical cord blood. We acknowledge that analyses on buffy coat have lower bio-variability and higher DNA yields (79). Fourth, although TL (80) and mtDNAc is known to be heterogeneous across blood cell types (81), we were unable to assess blood cell counts (e.g., peripheral blood mononuclear cells, hematopoietic cells, thrombocytes, lymphocytes, neutrophils). Thus, we are unable to eliminate differential blood compositions between prenatal control and intervention arms. Fifth, qPCR leads to an estimate of an individual’s relative TL, rather than the absolute number of base pairs (79), which limits the comparability of our findings to other RCTs to the directions of effects. Sixth, our qPCR assay fails to distinguish functional from dysfunctional mitochondria, as well as mtDNA retained within intact organelles, rather than mtDNA that may have leaked into cytoplasm due to nutrient-stress induced damage (82). Lastly, the average TL does not provide insights into telomere integrity, dysfunctionality, or the amount of short or critically short telomeres, which might relate to specific telomere pathologies and diseases (83). In conclusion, our results do not indicate that the provision of daily MMN-fortified BEP supplements to pregnant females affects newborn TL or mtDNAc. Future randomized interventions should assess the effects of preconception (84) and prenatal nutritional supplementation on absolute TLs and more granular measures of mitochondrial bio-energetic functioning (e.g., oxygen consumption rate, mtDNA mutations and deletions), while larger observational studies might further explore the TLs and mtDNAc of infants born with adverse phenotypes.

## Data Availability

All data produced in the present study are available upon reasonable request to the authors.

## Ethics

The study protocol was approved by the ethics committee of Ghent University Hospital in Belgium (B670201734334) and the ethics committee of Institut de Recherche en Sciences de la Santé (50-2020/CEIRES). An independent Data and Safety Monitoring Board (DSMB), comprising an endocrinologist, two pediatricians, a gynecologist, and an ethicist of both Belgian and Burkinabe nationalities, was established prior to the start of the efficacy trial. The DSMB managed remote safety reviews for adverse and serious events at nine and 20 months after the start of enrolment. The MISAME-III trial was registered on *ClinicalTrials.gov* (identifier: NCT03533712).

## Data availability

The informed consent form does not allow sharing of personal data outside the research team. Requests to access data must be directed to the ethics committee of Ghent University Hospital through. Supporting study documents, including the study protocol and questionnaires, are publicly available on the study’s website: https://misame3.ugent.be.

## Acknowledgements

The authors thank all the females from Boni, Dohoun, Karaba, Dougoumato II, Koumbia and Kari who participated in the study and the data collection team. We thank Nutriset (France) for donating the BEP supplements. The authors’ responsibilities were as follows—PK, CL, LH, LCT, TD-C, GH-C, YBM, MDB and SDS: designed the study; TD-C, YBM, LCT, and LO: conducted the research; GH-C: analyzed data and performed statistical analysis; DM, YBM, and TN conducted laboratory analyses; GH-C: developed the first draft and revised the manuscript; YBM, DM, TD-C, LT, BdK, LO, AA, KT, PK, LH, TN, SDS, MDB, and CL: critically reviewed the manuscript; and all authors: read and approved the final manuscript.

---

**Supplemental Figure 1.**
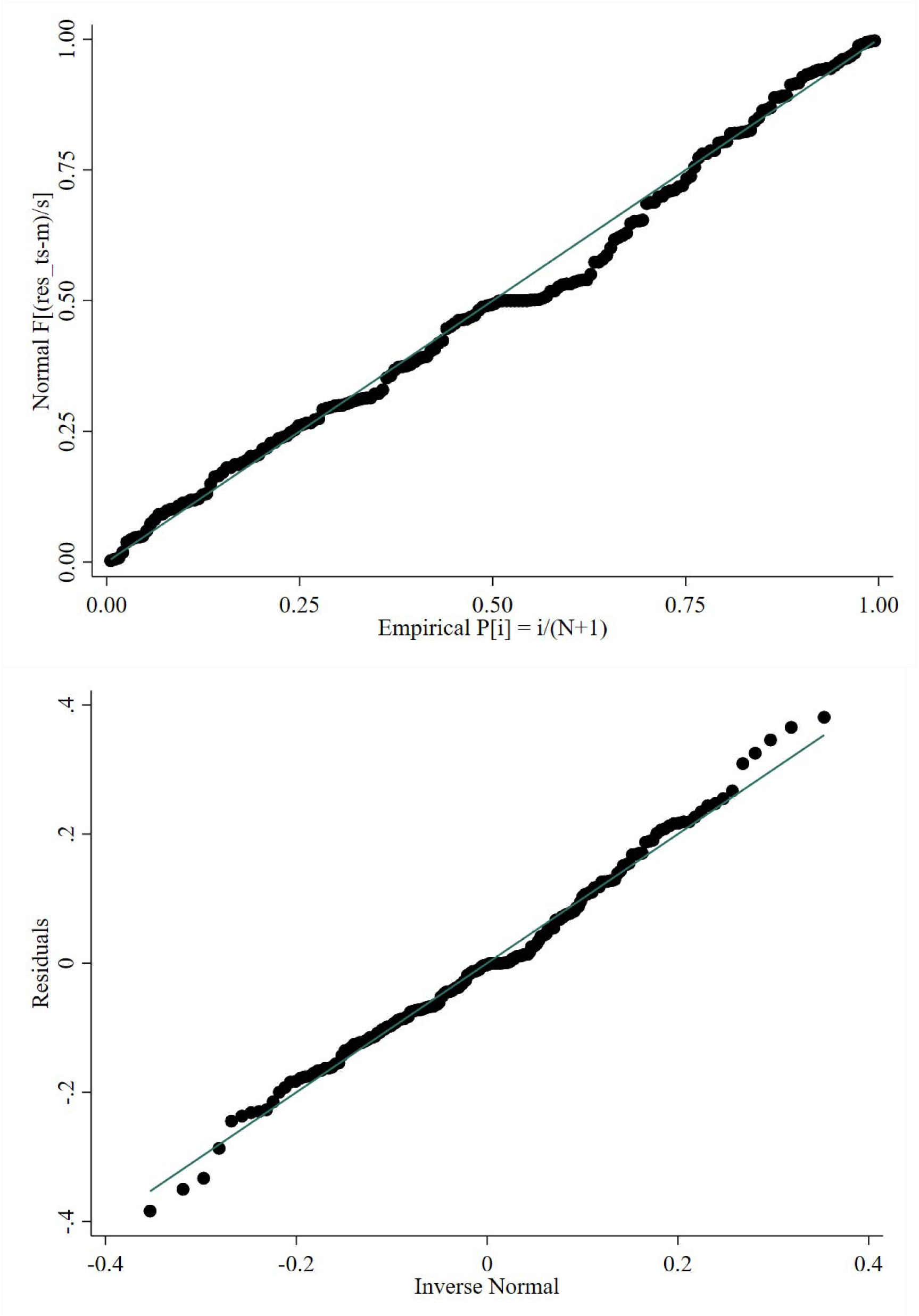
Standardized normal probability (top) and quantile-quantile (bottom) plots of unadjusted regression residuals for T/S ratio. T/S, telomere copy number to a single-copy gene number.

**Supplemental Figure 2.**
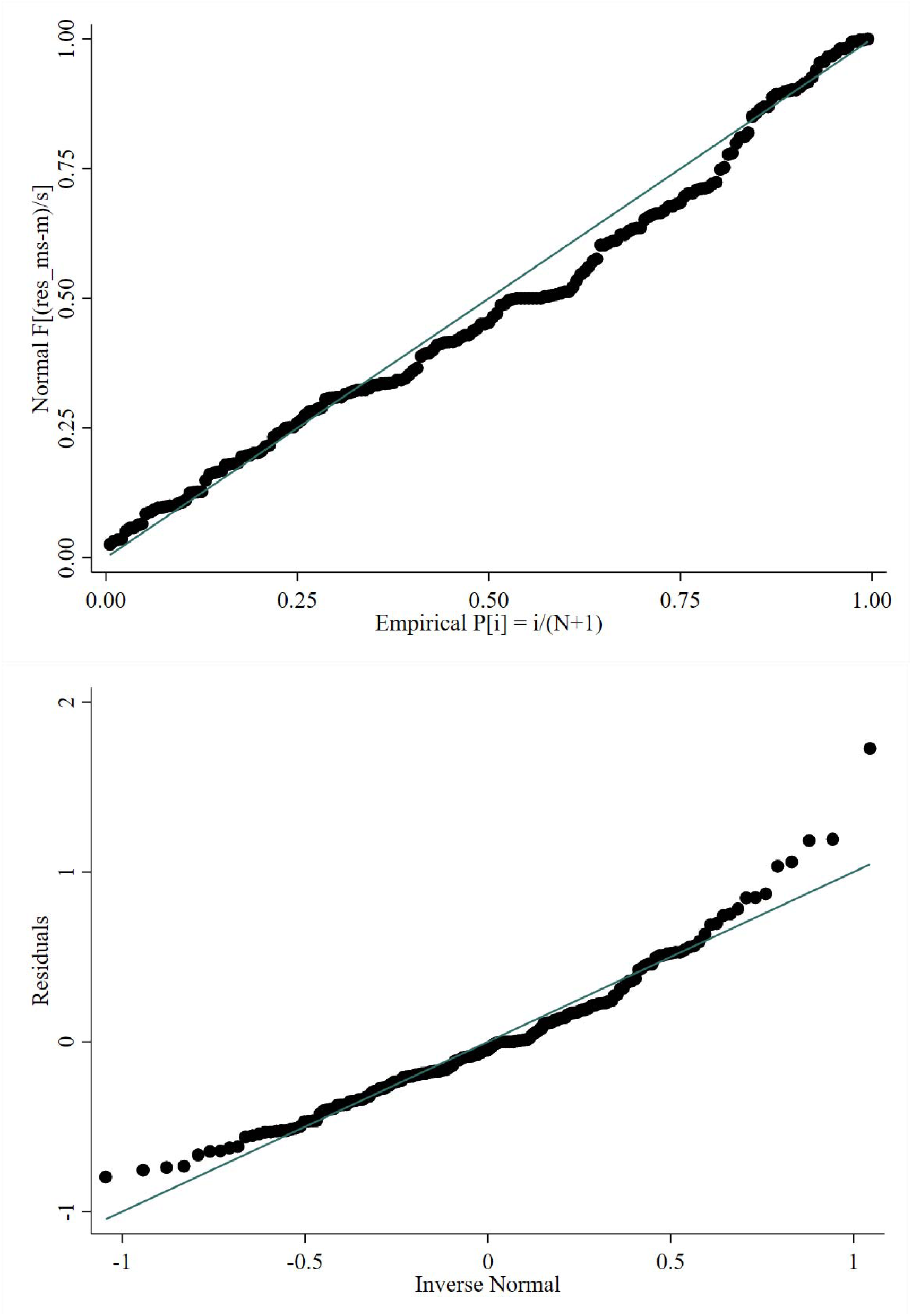
Standardized normal probability (top) and quantile-quantile (bottom) plots of unadjusted regression residuals for M/S ratio. M/S, mitochondrial gene copy number to a single-copy gene number.

**Supplemental Table 1.** Nutritional values of the ready-to-use supplementary food for pregnant and lactating women.

|  | Mean for 72g (serving size) |
| --- | --- |
| Total energy (kcal) | 393 |
| Lipids (g) | 26 |
| Linoleic acid (g) | 3.9 |
| α-Linoleic acid (g) | 1.3 |
| Proteins (g) | 14.5 |
| Carbohydrates (g) | 23.3 |
| Calcium (mg) | 500 |
| Copper (mg) | 1.3 |
| Phosphorus (mg) | 418 |
| Iodine (μg) | 250 |
| Iron (mg) | 22 |
| Selenium (μg) | 65 |
| Manganese (mg) | 2.1 |
| Magnesium (mg) | 73 |
| Potassium (mg) | 562 |
| Zinc (mg) | 15 |
| Vitamin A (μg RE)<sup>2</sup> | 770 |
| Thiamin (mg) | 1.4 |
| Riboflavin (mg) | 1.4 |
| Niacin (mg) | 15 |
| Vitamin B5 (mg) | 7 |
| Vitamin B6 (mg) | 1.9 |
| Folic acid (μg) | 400 |
| Vitamin B12 (μg) | 2.6 |
| Vitamin C (mg) | 100 |
| Vitamin D (μg cholecalciferol)<sup>3</sup> | 15 |
| Vitamin E (mg α-tocopherol)<sup>4</sup> | 18 |
| Vitamin K (μg) | 72 |

<sup>1</sup>Ingredients: vegetable oils (rapeseed, palm, and soy in varying proportions), defatted soy flour, skimmed milk powder, peanuts, sugar, maltodextrin, soy protein isolate, vitamin and mineral complex, stabilizer (fully hydrogenated vegetable fat, mono-, and diglycerides). IU, international unit, RE, retinol equivalent.
<sup>2</sup>1 μg vitamin A RE = 3.333 IU vitamin A.
<sup>3</sup>1 μg cholecalciferol = 40 IU vitamin D.
<sup>4</sup>1 mg $\alpha$ -tocopherol = 2,22 IU vitamin E.

**Supplemental table 2.** Differences in baseline characteristics of study participants, by MISAME-III trial arm.

| Characteristics | Control (*n* = 104) | Intervention (*n* = 90) | *P*-value<sup>5</sup> |
| --- | --- | --- | --- |
| **Health centre catchment area** |  |  | 0.88 |
| Boni | 17 (16.3) | 16 (17.8) |  |
| Dohoun | 14 (13.5) | 13 (14.4) |  |
| Dougoumato II | 13 (12.5) | 14 (15.6) |  |
| Karaba | 11 (10.6) | 8 (8.89) |  |
| Kari | 21 (20.2) | 21 (23.3) |  |
| Koumbia | 28 (26.9) | 18 (20.0) |  |
| **Household level** |  |  |  |
| Wealth index, 0-10 points | 4.32 ± 1.72 | 5.02 ± 1.77 | 0.006 |
| Household food insecurity<sup>2</sup> | 72 (69.2) | 64 (71.1) | 0.68 |
| Improved primary water source<sup>3</sup> | 62 (59.6) | 52 (57.8) | 0.80 |
| Improved sanitation facility<sup>4</sup> | 68 (65.4) | 54 (60.0) | 0.44 |
| Household size | 5.85 ± 4.34 | 6.89 ± 4.74 | 0.11 |
| Polygamous households | 38 (36.5) | 40 (44.4) | 0.26 |
| **Head of household** |  |  |  |
| Age, years | 32.3 ± 8.83 | 32.2 ± 9.31 | 0.95 |
| Male | 103 (99.0) | 90 (100) | 0.35 |
| Completed primary education | 63 (60.6) | 55 (61.1) |  |
| **Maternal** |  |  |  |
| Age, years | 24.0 ± 5.47 | 24.0 ± 5.70 | 1.00 |
| Ethnic group |  |  | 0.59 |
| Bwaba | 57 (54.8) | 51 (56.7) |  |
| Mossi | 36 (34.6) | 30 (33.3) |  |
| Other | 11 (10.6) | 9 (10.0) |  |
| Religion |  |  | 0.92 |
| Muslim | 47 (45.2) | 41 (45.6) |  |
| Animist | 26 (25.0) | 22 (24.4) |  |
| Protestant | 23 (22.1) | 19 (21.1) |  |
| Catholic | 6 (5.77) | 6 (6.66) |  |
| No religion, no animist | 2 (1.92) | 2 (2.22) |  |
| Completed primary education | 53 (51.0) | 37 (41.1) | 0.17 |
| Weight, kg | 58.9 ± 10.7 | 58.1 ± 7.88 | 0.54 |
| Height, cm | 162 ± 5.83 | 163 ± 5.83 | 0.36 |
| BMI, kg/m<sup>2</sup> | 22.4 ± 3.54 | 21.9 ± 2.65 | 0.27 |
| <18.5 kg/m<sup>2</sup> | 8 (7.69) | 4 (4.44) | 0.35 |
| Mid-upper arm circumference, mm | 263 ± 31.9 | 261 ± 24.0 | 0.56 |
| Subscapular skinfold, mm | 12.5 ± 7.18 | 11.8 ± 5.55 | 0.43 |
| Tricipital skinfold, mm | 11.9 ± 5.53 | 11.5 ± 4.38 | 0.59 |
| Haemoglobin (Hb), g/dl | 11.8 ± 1.34 | 11.8 ± 1.39 | 0.83 |
| Anemia (Hb <11 g/dl) | 23 (22.1) | 28 (31.1) |  |
| Severe anemia (Hb <7g/dl) | 0 (0) | 0 (0) | 1.00 |
| Gestational age, weeks | 9.43 ± 2.96 | 9.78 ± 3.41 | 0.45 |
| Trimester of gestation |  |  | 0.93 |
| First | 86 (82.7) | 74 (82.2) |  |
| Second | 18 (17.3) | 16 (17.8) |  |
| Parity |  |  | 0.26 |
| 0 | 26 (25.0) | 26 (28.9) |  |
| 1-2 | 49 (47.1) | 32 (35.6) |  |
| ≥3 | 29 (27.9) | 32 (35.6) |  |

<sup>1</sup>Data are frequencies (%) or means ± standard deviation. BMI, body mass index; Hb, hemoglobin. MISAME-III, Micronutriments pour la Santé de la Mère et de l'Enfant study 3.
<sup>2</sup>Assessed using FANTA/USAID's Household Food Insecurity Access Scale (79). Missing for one female in the control group.
<sup>3</sup>Protected well, borehole, pipe or bottled water were considered improved water sources.
<sup>4</sup>Flush toilet connected to local sewage or septic tank, or pit latrine with slab and/or ventilation were considered improved sanitation facilities.
<sup>5</sup>Two-sided t-test for continuous variables and Pearson's chi-squared test for categorical variables.

**Supplemental Table 3.** Efficacy of prenatal multiple micronutients-fortified BEP supplementation on relative telomere length and mitochondrial DNA content, by child sex.

| Birth characteristics | Control<sup>1</sup><br>(*n* = 104) | Intervention<sup>1</sup><br>(*n* = 88) | Unadjusted $\Delta^3$<br>(95% CI) | *P*-value | Adjusted $\Delta^3$<br>(95% CI) | *P*-value |
| --- | --- | --- | --- | --- | --- | --- |
| T/S ratio |  |  |  | 0.445<sup>4</sup> |  | 0.414<sup>4</sup> |
| Female | 56 (53.8) | 36 (40.9) | -0.025 (-0.100, 0.050) | 0.506 | -0.038 (-0.125, 0.049) | 0.385 |
| Male | 48 (46.2) | 52 (59.1) | 0.001 (-0.071, 0.068) | 0.967 | 0.003 (-0.075, 0.080) | 0.946 |
| M/S ratio |  |  |  | 0.682<sup>4</sup> |  | 0.600<sup>d</sup> |
| Female | 55 (53.3)<sup>2</sup> | 35 (39.8) | -0.094 (-0.323, 0.134) | 0.412 | -0.022 (-0.275, 0.230) | 0.859 |
| Male | 48 (46.6)<sup>2</sup> | 53 (60.2) | -0.145 (-0.363, 0.073) | 0.188 | -0.128 (-0.365, 0.109) | 0.284 |

<sup>1</sup>Values are frequencies (%). CI, confidence interval; T/S, telomere copy number to a single-copy gene number; M/S, mitochondrial gene copy number to a single-copy gene number
<sup>2</sup>M/S ratio for 103 women in the control group.
<sup>3</sup>Unadjusted and adjusted group differences ( $\Delta$ ) were estimated by fitting linear regression models. All models contained health centre, randomization block, and quantitative polymerase chain reaction plate as fixed effects to account for clustering by the study design. Adjusted models additionally contained a set of potential prognostic factors of outcomes including wealth index, maternal age, primiparity, gestational age, height, mid-upper arm circumference, body mass index, and haemoglobin level at study enrolment.
<sup>4</sup>Statistical significance was set at $P < 0.10$ for interactions.

**Supplemental Table 4.** Relative telomere lengths and mitochondrial DNA content, by birth phenotype.

| Birth characteristics | SGA (<10<sup>th</sup> percentile)<sup>2</sup> |  | *P*-value<sup>3</sup> | LBW (<2,500g) |  | *P*-value<sup>3</sup> | Preterm delivery (<37 weeks) |  | *P*-value<sup>3</sup> |
| --- | --- | --- | --- | --- | --- | --- | --- | --- | --- |
|  | No (*n* = 148) | Yes (*n* = 44) |  | No (*n* = 177) | Yes (*n* = 15) |  | No (*n* = 189) | Yes (*n* = 3) |  |
| T/S ratio | 1.01 ± 0.165 | 1.01 ± 0.134 | 0.987 | 1.01 ± 0.161 | 1.01 ± 0.121 | 0.931 | 1.01 ± 0.159 | 0.932 ± 0.101 | 0.367 |
|  | No (*n* = 147) | Yes (*n* = 44) |  | No (*n* = 176) | Yes (*n* = 15) |  | No (*n* = 188) | Yes (*n* = 3) |  |
| M/S ratio | 1.07 ± 0.441 | 1.16 ± 0.463 | 0.241 | 1.08 ± 0.426 | 1.21 ± 0.642 | 0.303 | 1.09 ± 0.438 | 1.33 ± 0.942 | 0.349 |

<sup>1</sup>Values are means ± standard deviation. LBW, low birthweight, SGA, small-for-gestational age; T/S, telomere copy number to a single-copy gene number; M/S, mitochondrial gene copy number to a single-copy gene number.
<sup>2</sup>Defined as the proportion of newborns with a birthweight below the 10<sup>th</sup> percentile of the International Fetal and Newborn Growth Consortium for the 21<sup>st</sup> Century (INTERGROWTH-21st newborn size standards for a given gestational age at delivery (40).
<sup>3</sup>Two-sided Student's *t*-test.

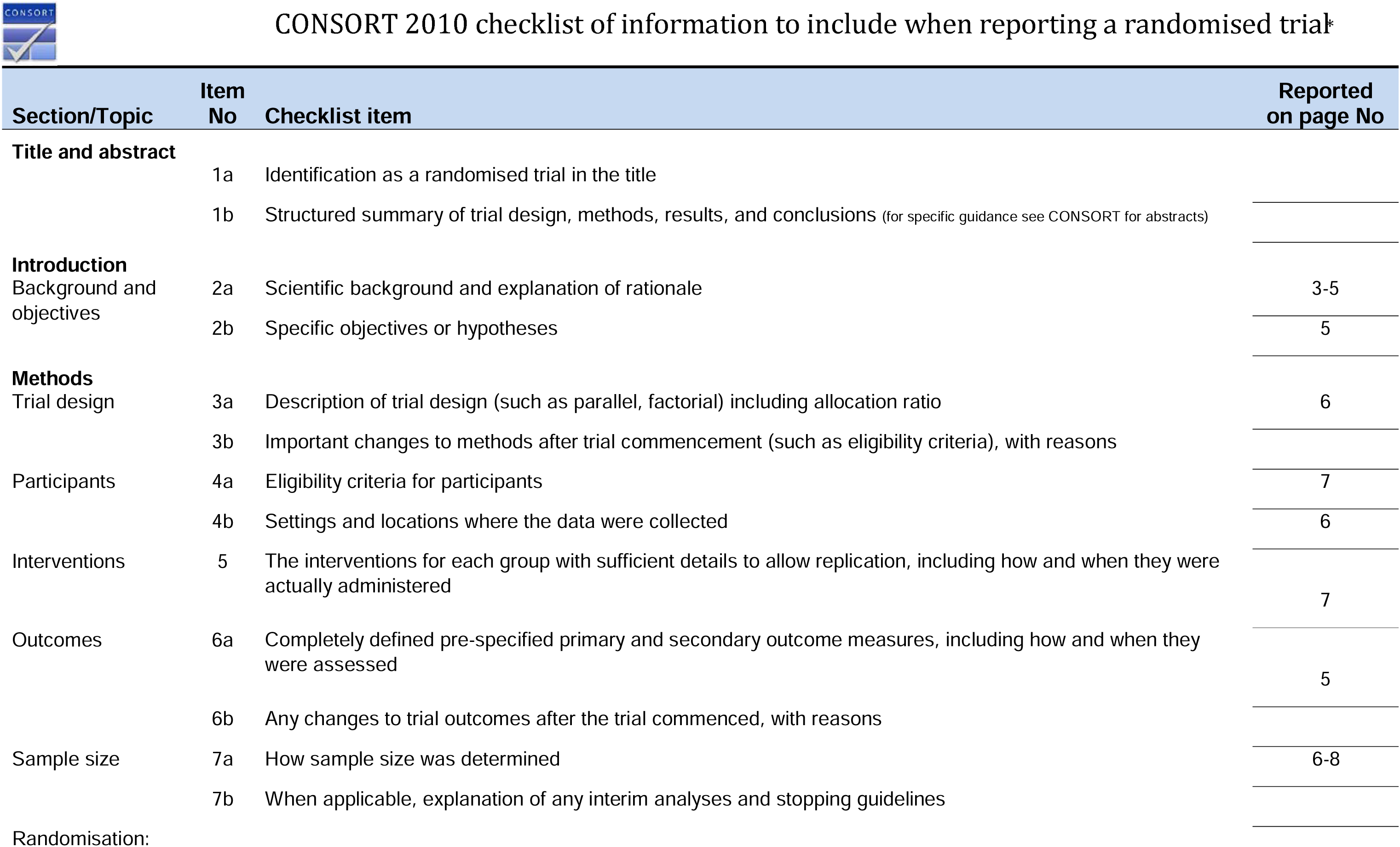
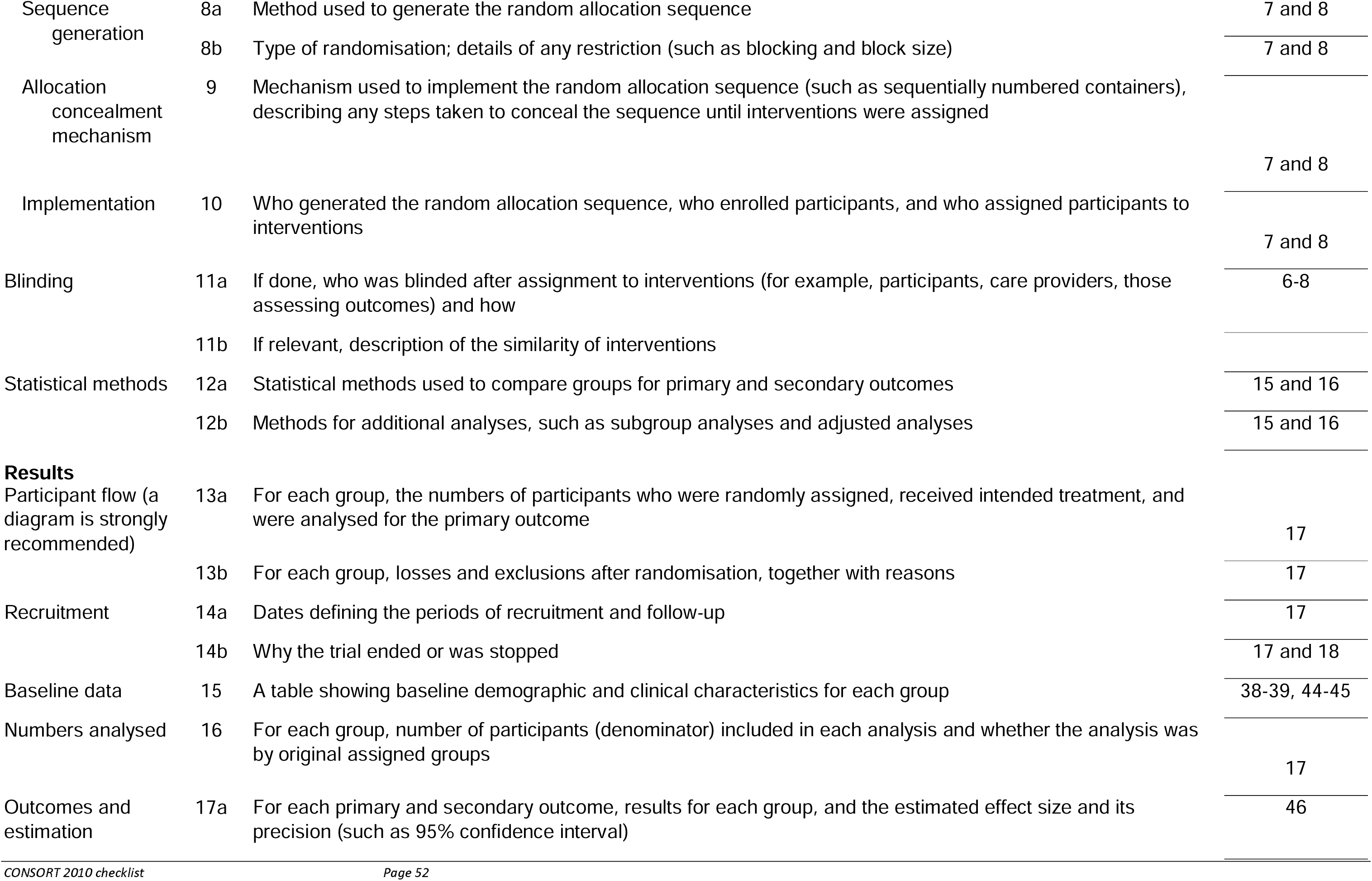
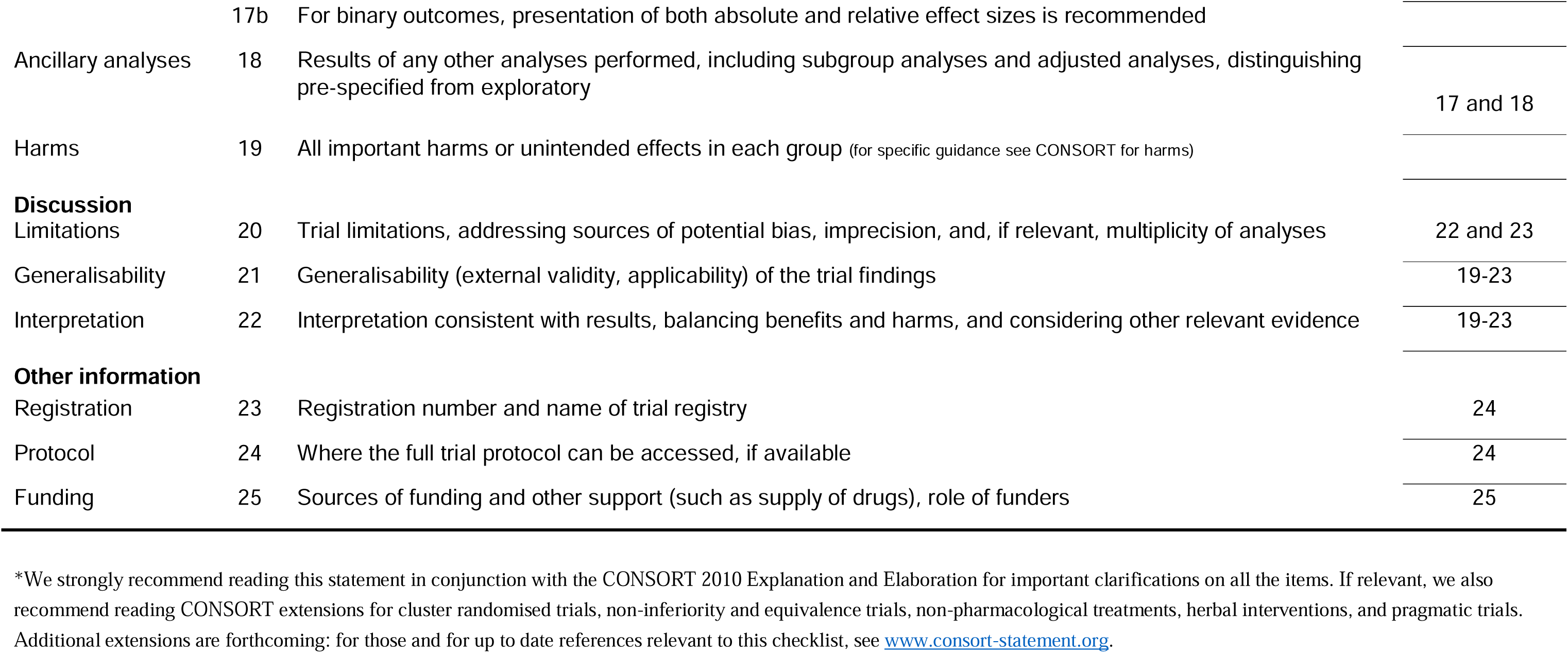

